# The waiting room: unmet sexual health service needs among men and gender-diverse individuals having sex with men in England, findings from an online, cross-sectional community survey in 2024

**DOI:** 10.1101/2025.10.01.25337058

**Authors:** Dana Ogaz, Dolores Mullen, George Baldry, Danielle Jayes, Dawn Phillips, Catherine M Lowndes, David Reid, Jordan Charlesworth, Erna Buitendam, David Phillips, Gwenda Hughes, Catherine H Mercer, John Saunders, Kate Folkard, Katy Sinka, Hamish Mohammed

## Abstract

**Background:** Sexual health service (SHS) delivery in England has shifted substantially following a rapid expansion of online services during the COVID-19 pandemic. While digital provision may improve reach, there are limited data on the extent of unmet need for in-person SHS in England. We sought to address this among men and gender-diverse individuals who have sex with men in England, a group disproportionately affected by sexual health inequalities.

**Methods:** We analysed data from “Reducing inequalities in Sexual Health” (RiiSH) 2024 (Nov/Dec 2024), an online cross-sectional survey of men and gender-diverse individuals who have sex with men residing in England. We assessed in-person SHS access and among those who tried to access a SHS in the last year - unmet need (i.e. participants who tried but were unable to access a SHS in-person) alongside reasons for inaccessibility. Using multivariable logistic regression, we examined sociodemographic and behavioural associations with unmet SHS need.

**Results:** Among 2,404 participants living in England (median age 45 years [interquartile range: 36-55], 88% White, 95% cisgender), 86% reported accessing in-person SHS ever, and 59% in the past year. Of those who tried to access in-person care in the past year, 12% (95% CI: 11%-14%) experienced unmet need, especially Outside London (15% vs. 8% in London). Common barriers included unavailable (50%) or inconvenient (41%) appointment times. In adjusted multivariable analysis, unmet need continued to be lower among participants living in London (aOR: 0.64 [95% CI: 0.44-0.92]), those financially comfortable (aOR: 0.69 [0.49-0.97]), and those reporting ≥1 marker(s) of sexual risk (e.g. HIV-PrEP use in the last year and/or in the last 3-4 months, the report of a bacterial STI diagnosis, engaging in chemsex, having had ≥10 male physical sex partners; aOR: 0.14 [0.10-0.20]). Unmet need was higher among participants with limiting long-term physical health conditions (aOR: 1.61 [1.12-2.30]) and those who reported ever using online postal self-sampling services for STI testing (OPSS) (aOR: 1.50 [1.07-2.09]).

**Conclusions:** Despite high SHS engagement, one in ten participants in this national community-based sample reported recent unmet need for in-person SHS. Equity-focused strategies are needed to meet evolving SHS demand. Local service delivery guided by joint strategic needs assessments could help address unmet need for SHS.

**Ethical considerations:** Ethical approval of this study was provided by the UKHSA Research and Ethics Governance Group (REGG; ref: R&D 524).

**Consent to participate:** Online informed consent was received from all participants and all methods were performed in accordance with guidelines and regulations set by the UKHSA REGG.

**Declaration of conflicting interest:** Authors have no conflicting interests to declare.

**Funding statement:** The RiiSH 2024 study and authors HM, JS, DR, CHM received partial funding support as part of The National Institute for Health Research Health Protection Research Unit in Blood Borne and Sexually Transmitted Infections at University College London in partnership with the UK Health Security Agency (https://bbsti.hpru.nihr.ac.uk). The funders had no role in study design, data collection and analysis, decision to publish, or preparation of the manuscript. All other authors received no specific funding for this work.

**Data availability:** The data that support the findings of this study are not publicly available to protect participant privacy. However, some aggregate data are available upon reasonable request from the UK Health Security Agency (UKHSA). Requests can be directed to.

## Background

In England, sexual health service delivery is primarily provided through publicly funded services which are free, open access (i.e. without referral from primary care), and confidential. Services include testing and treatment for sexually transmitted infections (STIs) and provision of STI and HIV prevention interventions (e.g. HIV-PrEP/PEP, doxyPEP, vaccination, condoms). Access is largely available through a network of specialist sexual health services (SHS) and increasingly via online platforms offering online postal self-sampling kits (OPSS) (i.e. self-collection of samples sent for laboratory testing with results sent to users). While some clinics offer walk-in services, many in-person appointments are dependent on online or telephone triage(1), with waiting times varying by local supply and demand.

Over the past decade, the sexual health landscape in England has undergone profound transformation(2). Service provision has been significantly impacted by the COVID-19 pandemic, accelerating already-evolving models of service delivery, amidst increasingly constrained public health budgets and clinical resources(3, 4). The proportion of OPSS testing provided through SHS has rapidly increased, representing 13% of all STI testing in 2019 (pre-COVID-19) to 42% in 2024(5). While OPSS has increased the reach of SHS for many(6-8), significant barriers remain. These include digital exclusion, low health literacy, and difficulties with self-sampling (particularly for blood samples), as well as concerns about confidentiality(9, 10). Service limitations exist on the provision of self-sampling services, such as locally imposed restrictions on eligibility and number of test kits available(11, 12). Inherent limitations of online interactions(13, 14) that include reduced ability to identify those with increased risk or complex needs, inability to deliver injections (e.g. vaccinations), and obscuring the need for clinicians to provide care for acute and complex conditions(13) pose significant challenges to comprehensive sexual health service provision.

Previous assessments of unmet need for STI and HIV testing in England have considered the general population(4), or gay, bisexual, and other men who have sex with men (GBMSM)(15), and have largely been based on researcher perceived need and defined on behavioural proxies (e.g. based on clinical history). While important, these assessments do not characterise unmet needs among those attempting to access care or who may already be experiencing SHS access inequalities. Given the changing sexual health landscape, especially following periods of major disruption and service reconfigurations, there is a need to examine in-person SHS accessibility. We use data collected from a large, online community survey to characterise SHS access and unmet need among men and gender-diverse individuals having sex with men, which are key groups more likely to experience sexual health inequalities in England(5, 16, 17).

## Methods

### Data collection and recruitment

The ‘Reducing inequalities in Sexual Health’ (RiiSH) 2024 survey is part of a series of yearly, online cross-sectional surveys, assessing the sexual health and well-being of a community sample of men and gender-diverse individuals who have sex with men in the UK. Recruitment in 2024 took place from 18^th^ November-11^th^ December. Survey recruitment was conducted through advertisements on social networking sites (Facebook, Instagram) and geospatial dating platforms (Grindr, Scruff, Jack’d and Recon). Survey methods have been previously reported(18, 19). In brief, participants eligible to take part included self-identifying men (cisgender or transgender), transgender women or gender-diverse individuals who were assigned male at birth, aged ≥16 years, resident in the UK and reporting sex with a man (cisgender and/or transgender) in the last year. Given differences in the commissioning of sexual healthcare across the four nations of the UK, analyses were restricted to those living in England.

### SHS access

We conducted descriptive analyses to examine in-person (i.e. face-to-face) SHS access (‘never’, ‘in the last year’). Among those with an in-person visit in the last year, we examined reasons for last visit (e.g. for STI testing) and why services were chosen (e.g. ‘I felt comfortable here’) (see Appendix I for question excerpts). Results were stratified by region of residence (London, Outside London), where we hypothesised greater accessibility in London over other areas. While we posited greater variation in urban vs rural areas, these examinations were not possible given lack of granularity in region of residence measures. We also report the proportion who reported that they had ever used OPSS for STI testing, defined as testing via private or public self-sampling services.

### SHS unmet need

Unmet need was defined as those unable to access an in-person SHS among those who tried. We report the proportion of participants with unmet need (%, 95% CI) and reasons for SHS inaccessibility by region of residence (London, Outside London).

### Factors associated with SHS unmet need

We examined factors associated with unmet need using binary logistic regression among those who tried to access a SHS in the last year. We present bivariate and multivariable associations and consider evidence of association where p<0.05. All specified covariates were included in multivariable modelling based on *a priori* consideration (age-group, sexual orientation) and/or associations in previous literature (e.g. disability, sexual risk)(20-22). These also included: financial stability (prioritised as a measure of deprivation given strong association with poor sexual health(5) relative to other available measures, including education); markers of sexual risk (see definition below) as a composite measure given the strong correlation between measures of sexual risk behaviours; as well as region of residence (dichotomised as London, Outside London given measure limitations, see above). We considered inclusion of self-report of physical conditions or illnesses lasting or expected to last for 12 months or more but prioritised the report of physical limitations due to these conditions (see definition below) as these could influence in-person accessibility. As an indirect measure of digital literacy, and/or of structural barriers to in-person SHS access, we also included the report of ever having used OPSS for STI testing.

Having markers of sexual risk was defined as those reporting HIV-PrEP use in the last year and/or in the last 3-4 months: the report of a bacterial STI diagnosis, engaging in chemsex (those that had used crystal meth, mephedrone or GHB/GBL), having had ≥10 male physical sex partners, and meeting partners through sex-on-premises venues, public sex environments (i.e. cruising environments), or at private sex parties (herein collectively called ‘venue risk’).

For those reporting a limitation associated with a long-term physical health condition, we created a binary measure (i.e. response of ‘Yes – a little’, ‘Yes, a lot’ vs ‘Not at all’ to the question: “Does your condition or illness reduce your ability to carry out day-to-day activities). Those not reporting a long-term health condition were classified as having no limitations.

Survey data was collected via the Snap Surveys platform (www.snapsurveys.com). Data management and analyses were conducted using Stata v17.0 or higher (StataCorp, College Station, TX, USA).

### Ethics statement

Ethical approval for RiiSH 2023 was granted by the UKHSA Research and Ethics Governance Group (REGG; ref: R&D 524). Online consent was collected from participants and there was no incentive offered to participate.

## Results

Of 2,758 participants recruited to RiiSH 2024, 2,404 were resident in England (87%) and included in analyses (excluded participants included those living in Scotland [n=194], Wales [n=100], and Northern Ireland [n=60]) (Appendix II). The median age of included participants was 45 years (interquartile range: 36-55); most were of White ethnicity (88%), cisgender male (95%), degree-educated (60%) and employed (78%). Nearly one-third resided in London (31%) (Table 1).

**Table 1:** Sociodemographic, clinical and behavioural characteristics of 1) all participants and 2) those who tried to access a sexual health service in the last year.

|  | All participants | Those who tried to access a SHS in the last year¶ |
| --- | --- | --- |
| Total | 2,404 (100.0%) | 1,629 (100.0%) |
| Sociodemographic characteristics |  |  |
| Age at survey completion (years) | 45 (36-55) | 45 (36-54) |
| Mean age at survey completion (years) | 45.4 (13.2) | 45.2 (12.7) |
| Age-group (3 categories) |  |  |
| 16-29 | 290 (12.1%) | 168 (10.3%) |
| 30-44 | 875 (36.4%) | 640 (39.3%) |
| 45 and over | 1,239 (51.5%) | 821 (50.4%) |
| Ethnic group (all categories) |  |  |
| White | 2,104 (87.5%) | 1,398 (85.8%) |
| Black | 55 (2.3%) | 44 (2.7%) |
| Asian | 134 (5.6%) | 99 (6.1%) |
| Mixed or other | 111 (4.6%) | 87 (5.4%) |
| Ethnic group (2 categories) |  |  |
| All other ethnic groups | 300 (12.5%) | 231 (14.2%) |
| White | 2,104 (87.5%) | 1,398 (85.8%) |
| Gender identity and sex at birth |  |  |
| All other gender identity groups | 110 (4.6%) | 76 (4.7%) |
| Cisgender male | 2,294 (95.4%) | 1,553 (95.3%) |
| Sexual orientation |  |  |
| Gay/homosexual | 1,940 (80.7%) | 1,366 (83.9%) |
| Bisexual, straight/heterosexual, or another way‡ | 464 (19.3%) | 263 (16.1%) |
| Region of residence (England) |  |  |
| London | 748 (31.1%) | 570 (35.0%) |
| South East | 323 (13.4%) | 205 (12.6%) |
| South West | 194 (8.1%) | 111 (6.8%) |
| West Midlands | 162 (6.7%) | 100 (6.1%) |
| North West | 316 (13.1%) | 218 (13.4%) |
| North East | 89 (3.7%) | 61 (3.7%) |
| Yorkshire and Humber | 189 (7.9%) | 122 (7.5%) |
| East Midlands | 168 (7.0%) | 97 (6.0%) |
| East of England | 206 (8.6%) | 141 (8.7%) |
| England - unknown | 9 (0.4%) | 4 (0.2%) |
| UK born |  |  |
| No | 548 (22.8%) | 403 (24.7%) |
| Yes | 1,856 (77.2%) | 1,226 (75.3%) |
| Educational qualifications |  |  |
| Below degree-level | 968 (40.3%) | 611 (37.5%) |
| Degree-level or higher | 1,436 (59.7%) | 1,018 (62.5%) |
| Employment |  |  |
| Not employed | 539 (22.4%) | 349 (21.4%) |
| Current employment (full/part-time, self-employed) | 1,865 (77.6%) | 1,280 (78.6%) |
| Comfortable financial situation |  |  |
| No | 1,306 (54.3%) | 883 (54.2%) |
| Yes (top two quartiles)§ | 1,098 (45.7%) | 746 (45.8%) |
| Clinical and behavioural characteristics |  |  |
| HIV status |  |  |
| Negative/unknown | 2,140 (89.0%) | 1,408 (86.4%) |
| PLWHIV (tested positive) | 264 (11.0%) | 221 (13.6%) |
| HIV-PrEP use since Dec 2023 (in last year) |  |  |
| No (includes PLWHIV and those never reported use) | 1286 (53.5%) | 596 (36.6%) |
| Yes | 1,118 (46.5%) | 1,033 (63.4%) |
| Ever used STI antibiotic post-exposure prophylaxis for the prevention of STIs |  |  |
| No | 2,073 (86.2%) | 1,343 (82.4%) |
| Yes | 331 (13.8%) | 286 (17.6%) |
| Ever reported recreational drug use associated with chemsex |  |  |
| No | 2,044 (85.0%) | 1,335 (82.0%) |
| Yes | 360 (15.0%) | 294 (18.0%) |
| No. of male physical sex partners since Aug 2024 (in last 3-4 months) |  |  |
| No sex/only virtual sex | 215 (8.9%) | 104 (6.4%) |
| 1 | 274 (11.4%) | 121 (7.4%) |
| 2-4 | 715 (29.7%) | 431 (26.5%) |
| 5-9 | 519 (21.6%) | 394 (24.2%) |
| 10 or more | 681 (28.3%) | 579 (35.5%) |
| Whether any male physical sex partners were new since Aug 2024 (in last 3-4 months) |  |  |
| No new partners | 539 (22.4%) | 258 (15.8%) |
| 1 or more new partners | 1,865 (77.6%) | 1,371 (84.2%) |
| Whether had vaginal/anal sex with a woman since Aug 2024 (in last 3-4 months) |  |  |
| No | 2,266 (94.3%) | 1,561 (95.8%) |
| Yes | 138 (5.7%) | 68 (4.2%) |
| Venue risk¶ |  |  |
| No | 1,595 (66.3%) | 968 (59.4%) |
| Yes | 809 (33.7%) | 661 (40.6%) |
| Bacterial STI diagnosis since Aug 2024 (in last 3-4 months) |  |  |
| No | 2,160 (89.9%) | 1,393 (85.5%) |
| Yes | 244 (10.1%) | 236 (14.5%) |
| Limiting long-term physical health condition |  |  |
| No | 1,904 (79.2%) | 1,290 (79.2%) |
| Yes | 500 (20.8%) | 339 (20.8%) |
| Limiting long-term mental health condition |  |  |
| No | 1,816 (75.5%) | 1,214 (74.5%) |
| Yes | 588 (24.5%) | 415 (25.5%) |
| Markers of sexual risk (composite) ‡‡ |  |  |
| No | 836 (34.8%) | 319 (19.6%) |
| Yes | 1,568 (65.2%) | 1,310 (80.4%) |
| Ever used a private or public OPSS§§ |  |  |
| No | 1,460 (60.7%) | 956 (58.7%) |
| Yes | 944 (39.3%) | 673 (41.3%) |
| Ever visited an in-person SHS |  |  |
| No | 330 (13.7%) | 46 (2.8%) |
| Yes | 2,074 (86.3%) | 1,583 (97.2%) |
| Tried to access in-person SHS in last year |  |  |
| No | 775 (32.2%) | ... |
| Yes, tried but unsuccessful | 202 (8.4%) | 202 (12.4%) |
| Yes, visited in-person SHS | 1,427 (59.4%) | 1,427 (87.6%) |

‡Includes those identifying as bisexual, straight, or another way. §Top two quartiles ("I am comfortable"/"I am very comfortable" from the question, "How would you best describe your current financial situation". ¶Met male partners through sex-on-premises venues, public sex environments (i.e. cruising environments), or at private sex parties. ‡‡ Includes reporting of: HIV-PrEP use in the last year (i.e. since Dec 2023), and/or in the last 3-4 months (i.e. since Aug 2024), the report of a bacterial STI diagnosis, engaging in chemsex, having had $\geq 10$ male physical sex partners, and meeting partners through sex-on-premises venues, public sex environments (i.e. cruising environments), or at private sex parties. §§ Online and postal self-sampling services for STI testing. ¶¶ Includes 4 participants with missing data on region of England residence and were not included in modelling (see Table 2). PLWHIV=people living with HIV. HIV-PrEP=HIV pre-exposure prophylaxis. STI=sexually transmitted infection. SHS=sexual health service. OPSS=Online and postal self-sampling for STI testing. RiiSH='Reducing inequalities in Sexual Health'.

### SHS access

Of all participants, 86% (2,074/2,404) reported they had ever had an in-person SHS visit (59% [1,427/2,404] in the last year) and 40% (944/2,404) reported ever using a self-sampling service for STI testing. Among those who reported *never* visiting a SHS, most (71% 235/330) also reported they had never used OPSS for STI testing. Of those who *had* ever visited an in-person SHS, 59% (1,225/2,074) reported that they had never used an OPSS (Table 1).

#### Reasons for last in-person SHS visit

Among those who ever visited a SHS and had done so in the past year (69% 1,427/2,074), wanting an STI test or a general sexual health check-up (62% 889/1,427) and HIV-PrEP access (48% 684/1,427) were the most common reasons for visits (Appendix III). Only 11% of all participants reported having had symptoms as a reason for their last visit.

#### Reasons for choosing an in-person SHS

Considering reasons for choice of SHS, close proximity to or ease to travel from home (66% 938/1,427) was most commonly reported. Those living in London were less likely to report that their clinic choice was because it was close to home compared to those living Outside London (58% vs 70%) but were more likely to cite proximity to work (24% vs 17%) or because of the service’s reputation (37% vs 16%) as reasons. Excellent staff (50% 719/1,427), and services that suited needs (50% 719/1,427) were also commonly reported by all as reasons for choosing the last SHS attended (Appendix IV).

#### SHS unmet need

Among those who tried to access a SHS in the last year, 12% (95% CI: 11%-14%, 202/1,629) could not, with this proportion varying by whether the participant lived in London (8% [6%-11%] vs. Outside London, 15% [12%-17%]; Table 1).

#### Reasons for in-person SHS inaccessibility among those with unmet need

Appointment unavailability (50% 102/202) and inconvenient appointment times (41% 82/202) were the most common reasons for inaccessibility (Appendix V). A higher proportion of those reporting unmet need in London reported inconvenient appointment times (46% vs 39%), no appointment availability (56% vs 49%) and waiting too long for an appointment (33% vs 16%) compared to those living Outside London.

### Factors associated with SHS unmet need

All participants who tried to access a SHS in the past year were included in regression models; 4 participants who did not specify region of residence were excluded from analyses (Table 1). In bivariate analysis, we found a lower likelihood of unmet need amongst older age groups (uOR: 0.55 [0.35-0.86] aged ≥45 vs 16-29); those living in London (uOR: 0.54 [0.38-0.76]; those reporting financial comfort (uOR: 0.59 [0.43-0.80]); and those with at least one marker of sexual risk (uOR: 0.14 [0.10-0.19]). There was a higher likelihood of unmet need amongst those who were bisexual, straight, or described themselves in another way (uOR: 1.82 [1.28-2.59] vs gay/homosexual), had a long-term physical health condition that caused limitations in their everyday life (uOR: 2.01 [1.45-2.77]), and those who had ever used a OPSS for STI testing (uOR: 2.01 [1.45-2.77]) (Table 2).

**Table 2.** Sociodemographic, clinical and behavioural characteristics associated with unmet need among those trying to access a sexual health service in the last year.

|  | uOR (95% CI) | aOR (95% CI) ‡‡ | p-value |
| --- | --- | --- | --- |
| Sociodemographic characteristics |  |  |  |
| Age-group (3 categories) |  |  |  |
| 16-29 | 1.00 (base) | 1.00 (base) | 0.493 |
| 30-44 | 0.69 (0.44-1.10) | 1.11 (0.66-1.85) |  |
| 45 and over | 0.55 (0.35-0.86) | 0.89 (0.53-1.51) |  |
| Sexual orientation |  |  |  |
| Gay/homosexual | 1.00 (base) | 1.00 (base) | 0.057 |
| Bisexual, straight/heterosexual, or another way‡ | 1.82 (1.28-2.59) | 1.48 (1.00-2.20) |  |
| London resident |  |  |  |
| No | 1.00 (base) | 1.00 (base) | 0.014 |
| Yes | 0.54 (0.38-0.76)§ | 0.64 (0.44-0.92) |  |
| Comfortable financial situation |  |  |  |
| No | 1.00 (base) | 1.00 (base) | 0.031 |
| Yes (top two quartiles)§ | 0.59 (0.43-0.80) | 0.69 (0.49-0.97) |  |
| Clinical and behavioural characteristics |  |  |  |
| Markers of sexual risk (composite) ¶ |  |  |  |
| No | 1.00 (base) | 1.00 (base) | <0.001 |
| Yes | 0.14 (0.10-0.19) | 0.14 (0.10-0.20) |  |
| Limiting long-term physical health condition |  |  |  |
| No | 1.00 (base) | 1.00 (base) | 0.011 |
| Yes | 2.01 (1.45-2.77) | 1.61 (1.12-2.30) |  |
| Ever used a private or public OPSS§ |  |  |  |
| No | 1.00 (base) | 1.00 (base) | 0.019 |
| Yes | 1.06 (0.79-1.43) | 1.50 (1.07-2.09) |  |

‡Includes those identifying as bisexual, straight, or another way. §Top two quartiles ("I am comfortable"/"I am very comfortable" from the question, "How would you best describe your current financial situation". ¶ Includes reporting of: HIV-PrEP use in the last year (i.e. since Dec 2023), and/or in the last 3-4 months (i.e. since Aug 2024), the report of a bacterial STI diagnosis, engaging in chemsex, having had ≥10 male physical sex partners, and meeting partners through sex-on-premises venues, public sex environments (i.e. cruising environments), or at private sex parties. § Online and postal self-sampling services for STI testing. ‡‡Includes 1625 observations; excludes 4 participants that did not specify England region of residence. HIV-PrEP=HIV pre-exposure prophylaxis. STI=sexually transmitted infection. SHS=sexual health service. OPSS=online and postal self-sampling for STI testing. RiSH=Reducing inequalities in Sexual Health'. uOR=unadjusted odds ratio. aOR=adjusted odds ratio.

In a multivariable logistic regression adjusted for age and sexual orientation (*a priori*), as well as region of residence, self-reported financial comfort, markers of sexual risk, limiting long-term physical health conditions and use of OPSS for STI testing, we found no evidence of association by age-group (uOR: 0.89 [0.53-1.51] aged ≥45 vs 16-29) or sexual orientation (aOR: 1.48 [1.00-2.20]). With adjustment, a lower likelihood of unmet need remained amongst London residents (aOR: 0.64 [0.44-0.92]), those financially comfortable (aOR: 0.69 [0.49-0.97]) and those with markers of sexual risk (aOR: 0.14 [0.10-0.20]). Those with a limiting long-term physical health condition (aOR: 1.61 [1.12-2.30]), and those that had ever used an OPSS (aOR: 1.50 [1.07-2.09]) had a higher likelihood of unmet need.

## Discussion

Analysis of this large, online community survey provides important insights into patterns of SHS use and unmet need of men and gender-diverse individuals who have sex with men in England. While most participants had ever accessed in-person SHS, unmet need was evident among those who had recently attempted to access a SHS in-person. Findings emphasise the importance of assessing whether current service models, many of which originate from adaptations made to SHS delivery during the COVID-19 pandemic, adequately meet contemporary needs. Equitable access to SHS will be critical for successful implementation of preventative interventions such as doxyPEP and 4CMenB vaccination as well as ongoing HIV combination prevention strategies in England. Ensuring accessibility will be essential to realising the full potential of emerging and existing preventative tools. Continued evaluation of mixed delivery models (in-person, online) will be needed to maximise SHS effectiveness and reach, especially when addressing symptomatic versus preventative needs.

In-person SHS use was high among participants (86% ever), with a visit in the last year reported among 59% of participants, highlighting the continued demand for in-person care even amid the growing availability of remote SHS delivery such as self-sampling. Having symptoms did not appear to be a major driver of attending SHS in this sample; however, HIV-PrEP access (48%) and STI testing (62%) were both common reasons for seeking an in-person visit. Both reasons may reflect the high uptake of HIV-PrEP among GBMSM in England, and adherence to national recommendations for quarterly STI and HIV testing for individuals having condomless sex with new partners(23-25), reinforcing the integral role of SHS for delivering STI preventative care. Only 40% had ever used an OPSS for STI testing, and many who had attended in-person SHS had never used an OPSS, suggesting that online options may not yet be fully accessible or substitutive for all. Understanding the reasons for preferring in-person versus OPSS, including trust, convenience, or additional support, requires further exploration.

Though most RiiSH participants accessed an in-person SHS, 12% of those who tried to access in-person care in the past year reported unmet need, with this proportion significantly higher for those groups known to be more likely to experience health inequalities. For example, we found higher unmet need reported by a higher proportion of participants identifying as bisexual or heterosexual, indicating ongoing barriers in service design or engagement strategies, particularly in services targeted to self-identifying gay and bisexual men. These findings highlight the need for more inclusive programmes of targeted interventions and a stronger equity lens in SHS provision and commissioning (26, 27). Financial comfort, living in London, and having at least one marker of sexual risk were independently associated with lower likelihood of unmet SHS need, while unmet need was higher among those with a long-term limiting health condition and those who had previously used OPSS. These findings underscore the complexity of access, which is not just about service availability but, at an individual level, must also consider capacity and preference to use those services. Our findings echo those from quasi-representative surveys of the general population, which reported a higher likelihood of unmet need among individuals with a limiting long-term condition, a stark reflection of persistent barriers to SHS faced by people living with disabilities(21, 28).

Unmet need also varied substantially by whether participants lived in or Outside London, suggesting that in-person SHS may not equally serve or be equally available to people living in different parts of England. This may also reflect greater SHS availability in London relative to other locations in England, where, by region, London has one of the highest number of SHSs in England (42/247)(29). Appointment unavailability and inconvenient appointment times were commonly-reported barriers to in-person SHS access, but were reported more frequently by London residents, potentially reflecting system pressure in high-demand services(2, 30). A mixed methods assessment of the use of OPSS services in the UK found that in-person SHS case-mix complexity increased following the introduction of OPSS(31). While the use of online services and lower levels of unmet need among those with markers of sexual risk suggests redirections of clinical complexity that prioritise those with the greatest clinical need, the impacts of this displacement among other potential service users are largely unexplored. Even less is known about the role of SHS as a gateway to broader healthcare in England, including mental health services, vaccination, and substance misuse support(32, 33). The importance of these touchpoints should be a key consideration in digital health planning, especially as digital services could expand(34).

Overall, our findings suggest the need for the evaluation of SHS service models, which will require a calibrated balance between in-person and online services in order to meet the needs of men and gender-diverse individuals who have sex with men more broadly. While digital services may increase reach and service capacity(35, 36), they should not replace in-person care where it is essential (e.g. vaccination) or could be the gateway to wider healthcare provision for vulnerable or marginalised groups who could benefit from further or integrated services(37). Barriers to in-person access, where appropriate, must be minimised. Locally informed, equity-focused strategies are essential for addressing these gaps. We posit that joint strategic needs assessments(38) conducted at local level and used to inform SHS commissioning could act as key levers for targeted interventions that could minimise unmet need. These assessments must consider local demand, structural inequalities, and service capacity (39) to reduce unmet need among those most at risk of being underserved, including sexual minorities, people with disabilities, and those living in areas with fewer SHS, as identified in this analysis.

### Strengths and limitations

A strength of this study is that it reports data from a large, community-based sample of people living across England. However, participants likely overrepresent individuals already engaged in SHS, and may not reflect experiences of under-engaged groups. Self-reported data may also be subject to recall or social desirability bias. While we did capture area of participant residence, this may not be geographically representative. This study did not have the statistical power to consider regional differences, or rural and urban settings, where STI and HIV epidemiology and SHS accessibility varies(5). Alongside, we have no information on distance to in-person SHS, or on the role of SHS loyalty to specific services where reputation may affect service availability which was more common as a reason for choosing a service among London-based participants. It is unclear whether OPSS were used following in-person inaccessibility. We found a higher likelihood of unmet need among those who had ever used an OPSS. However, due to the cross-sectional nature of this study, it is unclear whether this reflects barriers to in-person services or is an indication of routine STI testing practices among participants, such as regular STI testing (including through OPSS) as part of HIV-PrEP care pathways(23).

We defined unmet need largely as an expressed need(40) among those who self-reported that they had tried and been unable to access a SHS in the last year. However, expressed need may not equate to actual or objective need. Our measure could have variable effects. Unmet need could be underestimated as it does not capture individuals affected by broader individual-level (e.g., lack of knowledge, low health literacy) and structural barriers (e.g. regional SHS and OPSS availability, stigma) that may prevent SHS engagement altogether. At the same time, the use of expressed need could have overestimated unmet need, as participants facing SHS inaccessibility would be more likely to recall and report difficulties.

Service access questions (e.g. reasons for visit, why services were chosen) were based on formative work and cognitive testing(18) that preceded the COVID-19 pandemic, where online services were less common; however major service provision changes were in place at that time (e.g. HIV-PrEP accessibility, quarterly STI and HIV testing recommendations). Notably, even though this sample likely represents a population highly engaged with SHS, unmet need was still present. This may imply that even greater access challenges could exist for groups with a lower perceived risk of STI/HIV acquisition (e.g. heterosexual women, older age groups) or those who may have other unique barriers to using online services.

## Conclusion

While engagement with SHS was high, unmet need was evident among this highly health literate sample. These findings highlight the challenges of maintaining quality in-person SHS while scaling accessible digital options. Addressing these needs will require equity-focused, locally curated strategies and services.

## Supporting information

Appendices I, II, III, IV, V

## Acknowledgements

The authors wish to thank all participants to RiiSH 2024 and Takudzwa Mukiwa (Terrence Higgins Trust) for contributions to survey implementation. Authors acknowledge the members of the National Institute for Health and Care Research Health Protection Research Unit (NIHR HPRU) in Blood Borne and Sexually Transmitted Infections (BBSTI) Steering Committee: Professor Caroline Sabin (HPRU Director), Dr John Saunders (UKHSA Lead), Professor Catherine H Mercer, Professor Gwenda Hughes, Dr Hamish Mohammed, Professor Greta Rait, Dr Ruth Simmons, Professor William Rosenberg, Dr Tamyo Mbisa, Professor Rosalind Raine, Dr Sema Mandal, Dr Rosamund Yu, Dr Samreen Ijaz, Dr Fabiana Lorencatto, Dr Rachel Hunter, Dr Kirsty Foster and Dr Mamooma Tahir.

