## Appendices I, II, III, IV, V for "The waiting room: unmet sexual health service needs among men and gender-diverse individuals having sex with men in England, findings from an online, cross-sectional community survey in 2024"

**Supplementary material**

### **Appendix I. RiiSH 2024 question excerpts**

**Excerpt from “About sexual health services” section**

**Q92. (Compulsory question)**

**Routing: Asked to all**

Have you **ever visited** a sexual health service (i.e. had a **face-to-face appointment**)?

- No
- Yes

**Q93. (Compulsory question)**

**Routing: Asked where Q92=Yes**

When was the **last** **time** you visited a sexual health service?

- Less than one year ago
- One to two years ago
- More than two years ago

**Q94. (Compulsory question)**

**Routing: Asked where Q93=Less than one year ago**

Thinking back to your **last** visit to a sexual health service, was it:

- Before August 2024
- Since the start of August 2024

**Q95. (Non-compulsory question)**

**Routing: Asked where Q92=Yes**

Why did you attend a sexual health service (**the last time if more than once**)? [*tick all that apply*]

- I wanted an STI test or a general sexual health check-up
- I had no symptoms, but I was worried I might have an STI or HIV
- I had symptoms
- A sexual partner had symptoms
- A sexual partner was diagnosed with an STI
- Treatment after a previous positive test
- Check-up after a previous positive test
- As follow-up to an online test
- Ongoing HIV care and treatment
- To get ***post***-exposure prophylaxis (PEP) to prevent HIV (i.e. taken **after** sex)
- To get ***pre***-exposure prophylaxis (PrEP) to prevent HIV (i.e. taken **before** sex)
- Following sexual assault
- Following domestic violence
- I was told to attend by my GP/family doctor or another healthcare professional
- I needed condoms
- I needed contraception (other than condoms)
- I needed a vaccination
- I couldn’t get an online testing kit
- For another reason

*******

**Excerpt from “About sexual health services” section**

**Q97. (Compulsory question)**

**Routing: Asked where Q92=No OR Q93=One to two years ago, More than two years ago**

Thinking back to **the last year**, have you tried to get a face-to-face appointment to a sexual health service?

- No, I did not try to get a face-to-face appointment
- Yes, I did try to get a face-to-face appointment

**Q98. (Compulsory question)**

**Routing: Asked where Q97=** **Yes, I did try to get a face-to-face appointment**

In the **last year**, why were you unable to get a face-to-face appointment at a sexual health service? [tick all that apply]

- Unsuitable opening hours
- No appointments at convenient times
- No appointment availability
- I had to wait too long to get an appointment
- I was directed to online services instead
- Difficulty travelling to the appointment
- Work commitments
- Family commitments
- Another reason(s)

**Q100. (Compulsory question)**

**Routing: Where Q93=** Less than one year ago

Thinking about your **most recent** face-to-face visit to a sexual health service, please select what were the most important factors in your decision to use this service? [*tick all that apply*]

- The clinic was close by or easy to get to from my home
- The clinic was close by or easy to get to from my workplace
- The services on offer suited my needs very well
- No appointment was needed
- The staff were excellent
- The clinic has a great reputation
- No-one I know would likely have seen me going there so my privacy is protected
- The clinic offers/offered particular services which were important to me
- It was easy to get an appointment
- I felt comfortable here
- I felt involved in decisions in my care
- Staff had time to discuss my needs
- Another reason <free text box>
- Prefer not to say

*******

**Excerpt from “Testing for sexually transmitted infections (STIs) other than HIV” section**

**Q60. (Non-compulsory question)**

**Routing: Asked where Q57=Yes (Q57 not shown, Ever tested for STIs]**

Where have you **tested** for STIs other than HIV? *[tick all that apply]*

- At a sexual health service
- I used a free online self-sampling service [I took my own sample and sent it off for the result]
- I used a self-testing kit [taking my own sample and finding out the result immediately]
- At my GP practice
- At a community HIV testing service (that is not in a hospital or clinic)
- At an HIV clinic
- I used a private online self-sampling service (I had to pay for it) [I took my own sample and sent it off for the result]
- At a private medical practice (i.e. I had to pay for the service)
- At a mobile medical unit
- Somewhere else

***

**Excerpt from “About your sexual satisfaction and personal well-being” section**

**Q115. (Non-compulsory question)**

**Routing: Asked to all**

Do you have any physical conditions or illnesses lasting or expected to last for **12 months or more**?

- Yes
- No

**Q115a. (Non-compulsory question)**

**Asked where Q115=Yes**

Does your condition or illness / do any of your conditions or illnesses reduce your ability to carry out day-to-day activities?

- Yes, a lot
- Yes, a little
- Not at all

### **Appendix II. RiiSH 2024 participant flowchart**

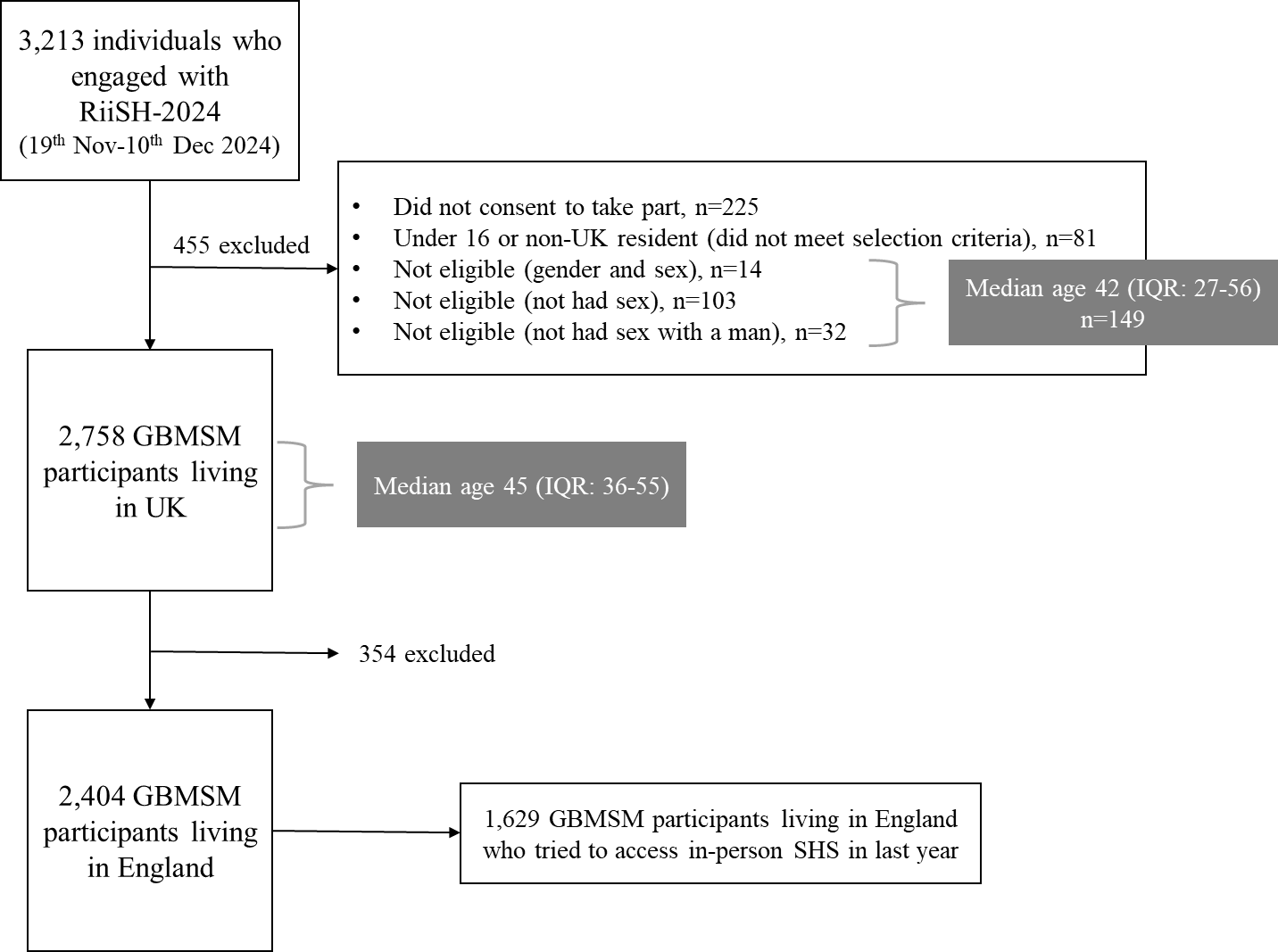

### **Appendix III: Reasons for last in-person SHS among those with a visit in the last year**

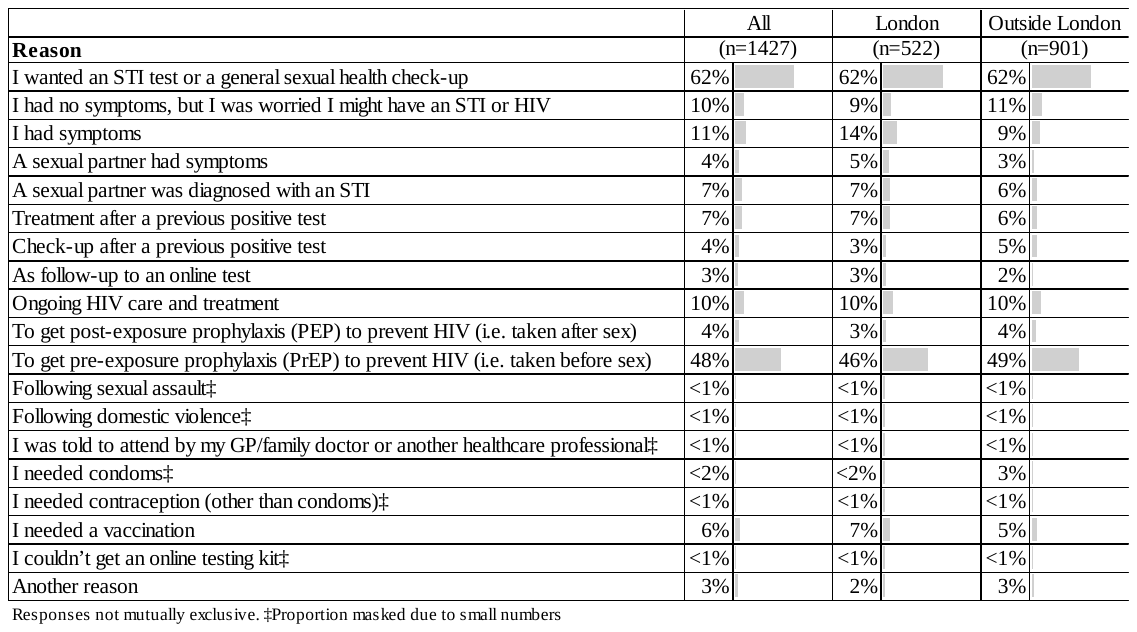

### **Appendix IV: Reasons for choosing last in-person SHS among those with a visit in the last year**

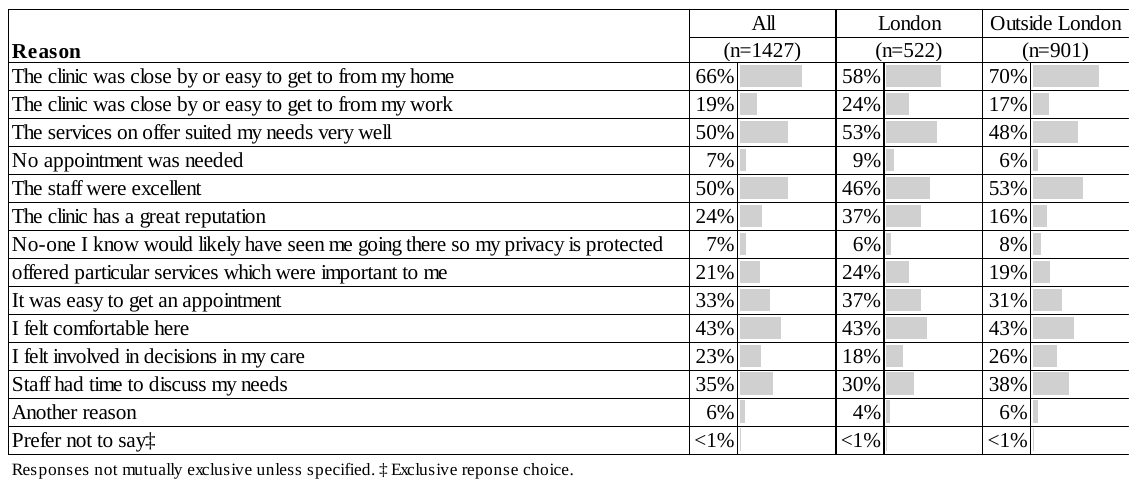

### **Appendix V: Reasons for in-person SHS inaccessibility among those who tried and were unable to access a SHS in the last year**

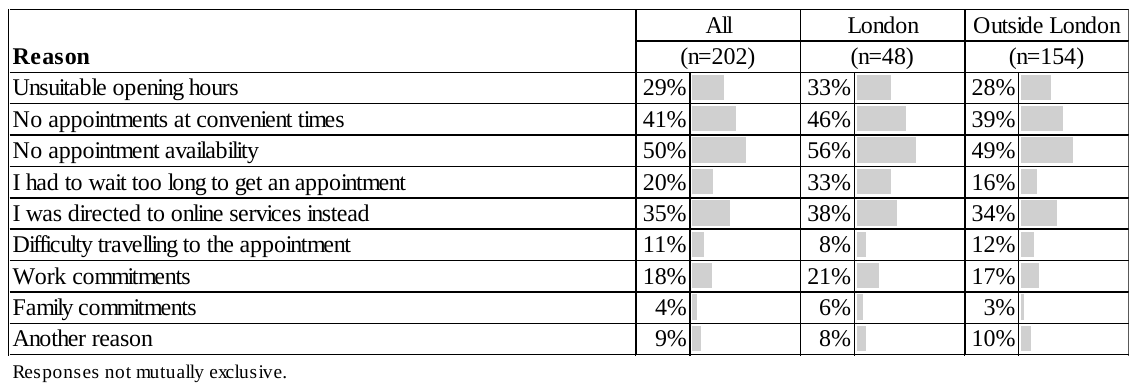
